# Integrating Wikipedia editing into health professions education: A curricular inventory and review of the literature

**DOI:** 10.1101/2020.03.19.20039339

**Authors:** Lauren A. Maggio, John M. Willinsky, Joseph A. Costello, Nadine A. Skinner, Paolo C. Martin, Jennifer E. Dawson

## Abstract

**Introduction:** Wikipedia is an online encyclopedia read by millions seeking medical information. To provide health professions students with skills to critically assess, edit, and improve Wikipedia’s medical content, a skillset aligned with evidence-based medicine (EBM), Wikipedia courses have been integrated into health professions schools’ curriculum. This study describes a literature review and curricular inventory of Wikipedia educational initiatives to provide an overview of current approaches and identify directions for future initiatives and research.

**Methods:** Five databases were searched for articles describing educational interventions to train health professional students to edit Wikipedia. Course dashboards, maintained by Wiki Education (WikiEdu), were searched for curricular materials. From these sources, key details were extracted and synthesized, including student and instructor type, course content, educational methods, and student outcomes.

**Results:** Six articles and 27 dashboards reported on courses offered between 2015-2019. Courses were predominantly offered to medical and nursing students. Instructors delivered content via videos, live lectures, and online interactive modules. Course content included logistics of Wikipedia editing, EBM skills, and health literacy. All courses included assignments requiring students to edit Wikipedia independently or in groups. Limited details of student evaluation were available.

**Discussion:** A small but growing number of schools are training HPE students to improve Wikipedia’s medical content. Course details are available on WikiEdu dashboards and, to a lesser extent, in peer-reviewed publications. There is limited evidence of the initiatives’ impacts on student learning, however, integrating Wikipedia into health professions education has potential to facilitate learning of EBM and communication skills, improve Wikipedia’s online content, and engage students with an autonomous environment while learning. Future considerations should include a thorough assessment of student learning and practices, a final review of student edits to ensure they follow Wikipedia’s Guidelines and are written in clear language, and improved sharing of teaching resources by instructors.

## INTRODUCTION

Wikipedia is an online encyclopedia consisting of over 6 million English-language entries, which is viewed more than 4.5 million times per hour, making it the seventh most visited site on the internet [1, 2]. While Wikipedia covers a wide range of topics, 30,000 English-language entries are dedicated to medical topics [2], which receive 10 million page-views per day across 286 languages [3]. Readers include patients and their families in search of information to prepare for health provider appointments or to understand information provided in such visits [4,5]. Many health professionals and students also consult Wikipedia, whether to provide patient care or study [6-9].

The nature of Wikipedia, which allows anyone to edit its entries, has raised concerns about health professionals and students using Wikipedia, non-medical professionals relying on it for medical information, and the potential impact on public health [3,10]. To address these concerns, health professions schools have launched courses to train learners how to critically assess and edit Wikipedia. For example, at the University of California, San Francisco, faculty offer medical and pharmacy students courses on improving Wikipedia’s quality [11,12]. At Queen’s University, all first-year medical students are required to take the course, *Critical Appraisal, Research and Lifelong Learning*, a major component of which trains students to critically assess and contribute to Wikipedia’s medical content [13]. The efforts of the Queen’s University students in 2017 amounted to more than 1,700 edits on 17 health topics, which were viewed over 3 million times in under a year.

At Queen’s University and other institutions, faculty have leveraged Wikipedia editing initiatives to support teaching evidence-based medicine (EBM) [13,14]. EBM is the “conscientious, explicit, and judicious use of current best evidence in making decisions about the care of individual patients” [15]. Most health professions schools are required to train students in EBM, which includes teaching learners to ask clinical questions, acquire and critically appraise information, and apply the information to a patient’s care based on clinical expertise and patient preferences [16,17]. Wiki Education (WikiEdu), a nonprofit started by the Wikimedia Foundation, provides support for Wikipedia educational initiatives across North America. To support initiatives across a variety of disciplines, WikiEdu provides online dashboards that facilitate course management, track editing progress and impact, and provide access to interactive training materials.

A recent scoping review focused broadly on situating Wikipedia as a health information resource [18]. While valuable, this review provides limited information on teaching approaches utilized, content covered, and evaluation techniques used in educational initiatives. Moreover, this review only includes educational initiatives described in the peer reviewed literature, which has been suggested as a narrow approach [19]. Thus, this study reviews online curricular materials and the relevant literature with an eye to provide educators with a detailed and holistic view of current approaches to using Wikipedia in HPE and to identify directions for future educational initiatives and research.

## METHODS

### Data Collection: Literature Review

On December 18, 2019, JC systematically searched MEDLINE via Ovid, Embase, CINAHL, ERIC, and Web of Science. Searches combined controlled vocabulary terms and keywords (See Appendix A for searches). Searches were limited to English. No date restrictions were applied. We also hand searched references of included articles. Citations were managed in EndNote.

For full-text review, we included articles describing curricular initiatives offered in health professions education (HPE), which were defined as programs leading towards careers in patient care. Articles were excluded if they described interventions using wiki software, but not specifically Wikipedia, and those using Wikipedia but not as part of a curriculum or course.

JC and NS independently reviewed article titles and abstracts. For articles that were unclear, LM served as a tiebreaker.

### Data Collection: Curricular inventory

On December 4, 2019, JC scanned WikiEdu’s dashboards to identify courses offered to HPE students [20]. A dashboard is a website provided to instructors by WikiEdu. Dashboards are designed to serve as an information and communication hub for faculty, students, and WikiEdu support staff engaged in courses. Dashboards, which instructors can tailor, provide tools to coordinate class assignments, track student edits, and serve as a platform to host course instructor and WikiEdu educational materials (Fig. 1).

**Figure 1:**
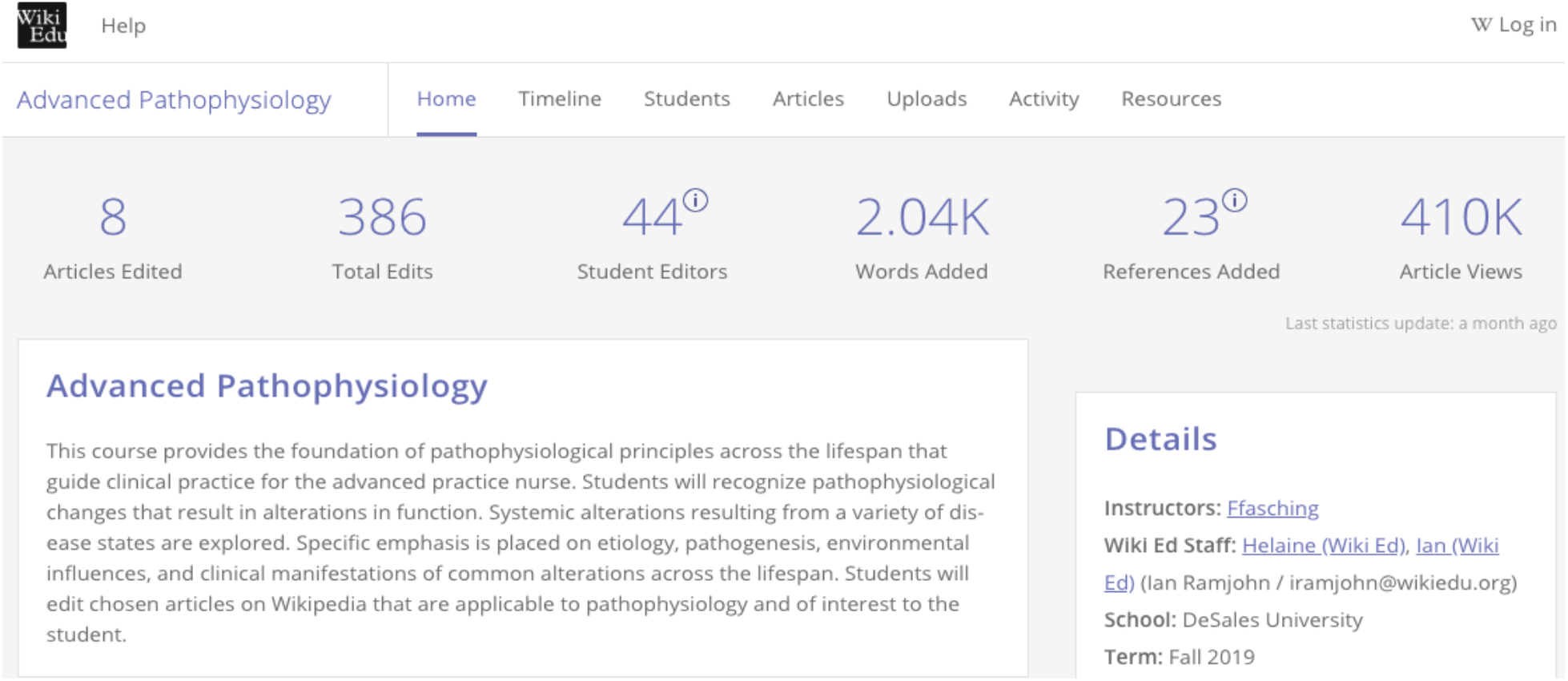
A screenshot of the WikiEd dashboard for the course *Advanced Pathophysiology* offered to 44 advanced practice nurses at DeSales University in Fall 2019 [30].

JC reviewed the all dashboard titles indexed as: *Communicating Science* and *Students in the Health Professions* to determine course audience. If the audience was unclear, he reviewed the dashboard’s content. If still unclear, the instructor, institution, and course name were googled to identify the course’s audience.

A dashboard was included if it reported that participating HPE students made at least one edit to a Wikipedia article. Dashboards also had to feature key course details, such as those found on the course timeline, to enable the extraction of relevant course data. We excluded dashboards that only used the dashboard’s tracking features (e.g., counts of student edits), but did not include curricular materials.

In the dashboard review, JC identified that courses were offered multiple times (e.g., a course offered in 2018 and 2019). In these cases, we included only the most recent dashboard, rationalizing that the latest iteration potentially reflected iterative lessons learned.

### Data Extraction

We designed a data extraction tool to capture details including, but not limited to, student and instructor type, course content, educational methods, and student outcomes. JC and LM independently extracted data from journal articles. Both JC and LM have taught medical students EBM and how to edit Wikipedia. JC and PM independently extracted dashboard data. PM has a background in education and community health and prevention research. LM, JC, and PM met via conference call to discuss and resolve coding differences.

### Synthesis and Analysis

We used Google Sheets to generate summary reports and descriptive statistics. We then collectively discussed similarities and differences in the initiatives and brainstormed implications for future educational initiatives and research.

## RESULTS

We included 27 dashboards [21-47] and six articles [11-14,48,49] (Fig. 2). Four articles [11, 12, 48, 49] described earlier accounts of six educational interventions also captured in our dashboard inventory such that there is considerable overlap between it and the literature review. For example, we include dashboard details from Azzam’s 2019 course and Azzam’s article that describes earlier iterations of the same course [11,23]. We first report dashboard results then article results.

**Figure 2:**
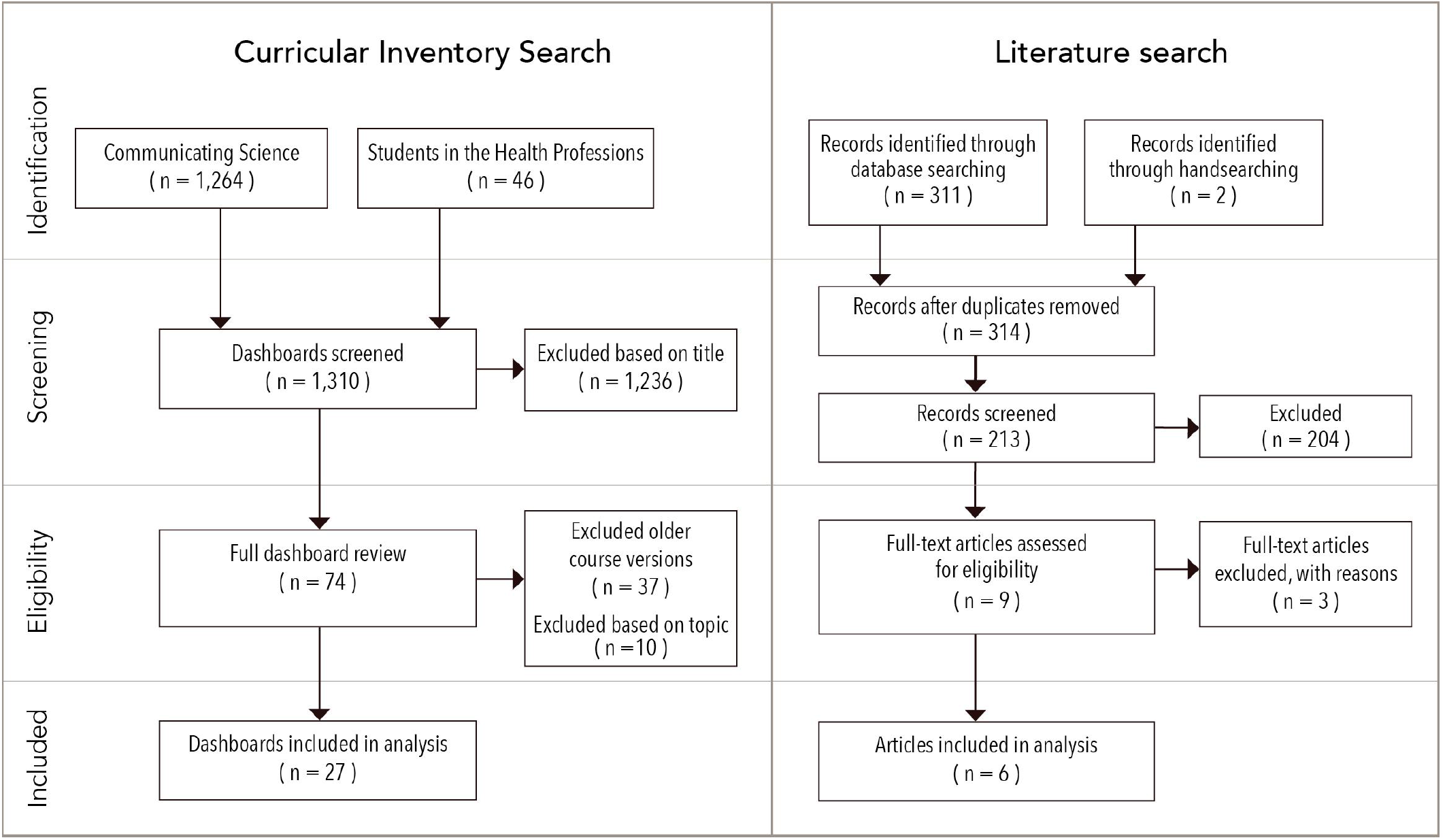
The search and selection process for curricular materials and literature search that describe educational interventions for health professions students that use Wikipedia.

### Dashboard Results

Courses were offered between 2015 and 2019, reflecting a growth in interest (Table 1). Courses ran from two [33] to 22 weeks [29], with an average of eight weeks. Courses were offered to medical (n=6) [23,29,31,32,34,44], nursing (n=4) [27,35,41,47], and audiology (n=2) students [40,45], among the leading areas that could be identified. Due to limited dashboard details, we were unable to determine the discipline in 12 courses, or if students were in the pre-clinical or clinical-phase of their studies. Course enrollment ranged from one [38] to 133 students [29].

**Table 1.** WikiEdu dashboard courses (N=27) by year in health professions education (HPE).

| Year | Courses | Students | Articles edited | # of edits | HPE areas |
| --- | --- | --- | --- | --- | --- |
| 2019 | 18 | 460 | 309 | 12,081 | Medicine, nursing, audiology, biomedical, translation |
| 2018 | 3 | 32 | 36 | 652 | Medicine, nursing, audiology |
| 2017 | 5 | 126 | 101 | 1,690 | Medicine, nursing, pharmacology, audiology, research |
| 2016 | 1 | 119 | 31 | 1,190 | Pharmacology |

#### Instructors

Instructors included physicians, librarians, nurses, and pharmacists. In two instances, students served as instructors [28,44]. While we limited our analysis to the most recent dashboard iterations, 16 instructors had previously offered courses. For example, Lebowitz offered a course in 2017, 2018, and 2019 [36]. All courses were supported by WikiEdu staff members, which was associated with the use of the dashboard.

#### Dashboard Focus

WikiEdu dashboards are intended to support courses for training students to edit Wikipedia’s health pages. However, several courses integrated Wikipedia assignments as an approach for students to apply their learning of medical topics. For example, the course, *Foundations of Clinical Trauma Psychology*, which focused on trauma disorders and treatments, incorporated a Wikipedia assignment to allow students to apply and broadly share their newly acquired knowledge [26]. In a course on medical-related linguistic translation, Wikipedia was specifically used to provide students opportunities to apply their medical translation skills and contribute to Wikipedia’s Spanish-language version [38]. In this regard, several instructors highlighted Wikipedia as a conduit for students’ work to have impact beyond the classroom. To facilitate this goal, three instructors addressed health literacy in their courses to ensure students’ Wikipedia edits would be appropriate for its public audience [23,24,28]. For example, one course contained the lecture, “Writing for the Public, Considerations About Health Literacy,” and provided learners a readability calculator and guide for avoiding medical jargon [24].

#### Content Delivery

Instructors delivered content through videos, live lectures, guest speakers, and WikiEdu modules. For example, four instructors incorporated guest lecturers with three being librarian-led [24,28,29,36]. All course dashboards included WikiEdu modules, which were primarily presented as text documents, but also included videos and quizzes. We identified 44 distinct WikiEdu modules across the dashboards, including those focused on the mechanics of editing Wikipedia, such as “Contributing Images and Media Files” (included in 25 courses) and “Editing and “Drafting in the Sandbox” (14), while other modules covered broader topics, such as “Peer Review” (21). (See Appendix B for included modules). All courses include the “Wikipedia Policies” module, which introduced the overall ethos of Wikipedia and its Five Pillars, which encourages editing from a neutral point of view and treating all editors with respect and civility [50].

Seventeen instructors customized their dashboards to orient students to the course. For example, five instructors supplemented WikiEdu modules in their orientation materials with video and audio files, which often featured the instructors themselves [23,24,28,31,36]. Four instructors required full-day, on-site orientation sessions [23,24,28,36].

Two dashboards specifically mentioned EBM [27,34]. However, through WikiEdu modules and additional instructor efforts, such as guest speakers and library resource guides, learners were trained in EBM skills, including to identify and articulate knowledge gaps (23 courses) and acquire (8) [23-25,28,31,32,34,36], appraise (26), and apply (26) information.

Four instructors in five courses linked or embedded additional resources in the dashboards, such as cloud apps (e.g., GoogleDocs) for document sharing [21-23,28,31], while five instructors utilized video conferencing tools (e.g., Zoom) to host synchronous online sessions and digital office hours [23,24,28,31,36]. One instructor linked to their institution’s learning management system [47]. All courses utilized the Wikipedia-specific “My Sandbox” feature, which is a tool offered by Wikipedia, that allows students to practice drafting edits in a simulated Wikipedia environment, prior to making live changes to Wikipedia. In addition to providing a low-stakes environment for practice editing, instructors encouraged or in some cases required students to request peer review of their edits while they were in their My Sandbox.

#### Wikipedia Assignments

All courses assigned students to edit a Wikipedia article required the application of EBM skills to identify gaps in a page’s content, acquire and appraise information to fill the gap, and then apply that information to make Wikipedia edits. Students primarily completed assignments independently. However, in nine courses students could work independently or in groups [21,22,27,32,36,41,42,45,47]. Four instructors had students work in groups [25,30,33,34] and in two courses we were unable to make such a determination [26,44]. In 12 courses, students self-selected their article to edit. To guide their choice, seven instructors assigned the WikiEdu modules “Finding Your Article” [21,23,28,31,38,39,44] and four assigned “Choose Your Article” [21,38,39,47]. Fourteen instructors required students to select articles based on the course topic and/or select from an instructor-provided list. A single instructor assigned students an article [25].

Students were also frequently assigned to peer review fellow students’ work (24 courses) and to respond to peer comments (21), which was supported by a WikiEdu module (21) and/or WikiEdu’s Peer Review Rubric [23,28,31,37,51]. Fourteen courses also required that students make final presentations describing their edits. Less common assignments included: reflective essays [30,43,39,27,32], Work-in-Progress reports [23,24,28,31,36,42], and requesting students nominate work for the “Did you Know?” feature on Wikipedia [22,42,45,46].

#### Evaluation

Beyond assignments, dashboards provided limited details on assessment of student learning. However, there was evidence of overall course evaluations and evaluations of each course’s impact on Wikipedia. For example, in five courses included end-of-course feedback sessions [23,24,28,31,36] and in one course students completed a final survey [32].

The 27 dashboards reflecting the work of 737 HPE students, indicated that 15,613 edits had been made on 477 articles, including the addition of 4,682 references and 42 images, which had been viewed 42.2 million times as of January 23, 2020.

### Literature Results

The six included articles covered eight courses – for which we provide a summary of general characteristics (Table 2) and educational approaches (Table 3) – with only two courses not featured in the dashboard inventory [13,14]. Courses were offered solely to medical students and pharmacy students, while one was interdisciplinary. Three authors reported multiple offerings of the course making it difficult to discern the number of enrolled students per offering [11,13,14]. For example, Badgett reported two course offerings with one cohort of students enrolled in 2007 and the other in 2008 for a total of 39 learners taught [14]. Course instructor identities were vague with authors noting those teaching simply as faculty or instructors without indication of profession except in the case of librarians, which were reported in four courses [11,48]. In Apollonio’s course near-peer students who had previously completed the course taught a hands-on demonstration session [12]. Five authors stated course objectives, which related to EBM and enhancing Wikipedia’s content (Table 3).

**Table 2:** Characteristics of educational initiatives (N=8) to teach Wikipedia editing to HPE students as reported in the literature.

| Author, year | Students (cohorts) | Audience | Level | Instructor type | Duration | Status | Setting |
| --- | --- | --- | --- | --- | --- | --- | --- |
| Apollonio, 2018 [12] | 119 | Pharmacy | Preclinical | Unspecified; near peer students | 9-weeks | Required | USA |
| Azzam, 2017 [11] | 43 (4) | Medical | Clinical | Instructors, librarians | 4-weeks | Elective | USA |
| Badgett, 2011 [14] | 39 (2) | Medical | Clinical | Unspecified | Unspecified | Elective | USA |
| Evenstein, 2017 [49] | 62 | Medical, dental, PhD, faculty, staff | Unspecified | Faculty | Semester | Elective | Israel |
| Joshi, 2019 [48] (MUSC)<sup>a</sup> | 50 (2) | Medical | Clinical | Faculty, librarians | 4-weeks | Elective | USA |
| Joshi, 2019 (UCF) | 12 | Medical | Clinical | Faculty, librarians | 4-weeks | Elective | USA |
| Joshi, 2019 (UCSF) | 16 (3) | Medical | Clinical | Faculty, librarians | 4-weeks | Elective | USA |
| Murray, 2020 [13] | 101 | Medical | Preclinical | Unspecified, Wikipedia medical editor | Unspecified | Required | Canada |

Note
<sup>a</sup> University of California, San Francisco (UCSF), University of Central Florida (UCF), and Medical University of South Carolina (MUSC)

**Table 3:** Course objectives provided by instructors in WikiEdu Dashboard for teaching Wikipedia editing to HPE students.

| Author | Course objectives |
| --- | --- |
| Apollonio, 2018 [12] | * Develop the professional skills associated with pharmacy practice by improving medicines information in Wikipedia. |
| Azzam, 2017 [11] | * Hone information retrieval and assessment skills;<br>* Practice communicating medical knowledge to an exceptionally broad global audience;<br>* Expand their sense of the health provider's roles in the Internet Age. |
| Badgett, 2011 [14] | * Learn components of evidence-based practice. |
| Evenstein, 2017 [49] | * Create quality medical-related content in Wikipedia;<br>* Be better consumers of online information;<br>* Become active members of the information culture by taking part in a collaborative construction of knowledge. |
| Murray, 2020 [13] | * Evaluate Wikipedia as a platform for sharing medical information;<br>* Search for and locate medical information;<br>* Enhance Wikipedia using EBM skills;<br>* Analyze claims of a Wikipedia article on complementary and alternative medicine. |

### Content delivery

All instructors blended in-person and online elements (e.g., hands-on skills labs, recorded lectures). For example, the three courses reported by Joshi used teleconference software to enable students to remotely attend weekly meetings [48]. Except for Badgett’s course, which was offered before WikiEdu’s founding, all courses utilized WikiEdu modules and dashboard. Evenstein’s course also described using the institution’s learning management system, Moodle, to organize course material. Of note, all courses focused on the English-language version of Wikipedia, except for Evenstein’s, which edited the Hebrew-language Wikipedia [49].

### Assignments

Instructors required students to complete a Wikipedia editing assignment (Table 4). Students were provided varying levels of autonomy in selecting a Wikipedia article to edit. For those that were prescriptive, article selection was commonly related to the course’s topic. As a component of the editing assignment, three courses required students to peer review each other’s edits [13,48,49]. We propose that by completing these assignments students had the opportunity to practice EBM skills.

**Table 4:** Details of educational activities, assignments and evaluation from courses to teach Wikipedia editing to HPE students.

| Author | Educational Activities | Assignments | Evaluation Methods | Evaluation findings |
| --- | --- | --- | --- | --- |
| Apollonio 2018 [12] | * Complete readings and Wiki Edu modules<br>* Attend in-person project overview and editing session | In small groups, edit instructor-selected Wikipedia pages | * Surveys at multiple timepoints in the course<br>* Analysis of Wikipedia edits | Student knowledge of Wikipedia editing processes increased and their edits improved quality of Wikipedia pages edited |
| Azzam, 2017 [11] | * Participate in in-person 2-day orientation<br>* Attend optional virtual instructor office hours | Independently edit Wikipedia page of student's choice | * Interviews<br>* Focus groups<br>* Analysis of Wikipedia edits | Students improved quality of Wikipedia pages edited, including readability, and credited the course in improving communication skills |
| Badgett, 2011 [14] | * Attend 4-hour in-person session<br>* Complete online training (unspecified) | Independently edit a Wikipedia page of student's choice | * Post-course survey<br>* Analysis of Wikipedia edits | Students were able to edit Wikipedia and were receptive to the course |
| Evenstein, 2017 [49] | * Attend in-person sessions<br>* Participate in online discussion forums on Moodle | * Independently write a Wikipedia page from an instructor-curated list<br>* Peer review classmate<br>* Final presentation | * Post-course survey<br>* Interviews<br>* Analysis of Wikipedia edits | Students rated their satisfaction with the course and their self-perceived knowledge as high |
| Joshi, 2019 [48]<br>MUSC<br>UCF<br>UCSF | * Attend in-person orientation session<br>* Complete Wiki Edu modules<br>* Attend weekly work-in-progress support sessions | * Independently or in teams edit a Wikipedia article of choice or from an instructor-curated list<br>* Peer review a classmate's work<br>* Final presentation | Post-course survey | Students reported increased perception of Wikipedia's reliability, an inclination to encourage peers to edit Wikipedia, improved ability to practice EBM, and increased desire to contribute to the public good |
| Murray, 2020 [13] | Complete Wiki Edu modules | * Independently and in small groups critique Wikipedia pages flagged for improvement<br>* Locate information to improve page<sup>b</sup>,<br>* Peer review another group's proposed edits. | * Post-course survey<br>* Analysis of student assignments | Students reported satisfaction with the experience and opportunity to contribute to Wikipedia, but also identified barriers related to mechanics of editing Wikipedia and difficulty engaging with Wikipedia community |

### Evaluation

Instructors utilized surveys, focus groups, interviews, and analysis of student Wikipedia edits to evaluate their students and courses (Table 4). Primarily surveys queried students’ satisfaction, but also self-perceived knowledge of Wikipedia and intentions to continue editing. Apollonio surveyed students at three points in the course using knowledge questions to objectively gauge increases in students’ knowledge of Wikipedia’s editing processes [12].

Students reported overall satisfaction with the courses and self-perceived improvements in their knowledge and skills. For example, the majority of students in the three courses described by Joshi self-reported increases in their ability to practice EBM [48]. Apollonio, based on pre-post surveys, concluded that her students’ knowledge of Wikipedia editing improved by course end [12]. Authors also analyzed students’ survey free-text responses. For example, by analyzing write-in comments, Murray identified course elements that the students appreciated and those identified as barriers, such as difficulties engaging with the broader Wikipedia editor community and students’ hesitancy to make live edits [13]. Additionally, Evenstein’s students described appreciating the opportunity to give back to the community via their edits, but also expressed that certain course elements (e.g., insufficient time dedicated to learning particular tasks) hindered the experience [49]. Interview and focus group data presented a similar mixed picture. Azzam’s students described difficulties in ensuring the readability of their edits and lamented that editing was more work than expected, but also highlighted that “it feels good to contribute to Wikipedia” and that they appreciated the potential global significance of their edits [11].

Four instructors analyzed the number of student edits made and references added or deleted to students’ to assess the quality and potential impact on Wikipedia [11-14]. Instructors also tracked the durability of student edits such that they were not reversed and used this as an indicator of quality edits. In two notable instances, the instructors went further. Azzam utilized natural language processing software to compute a “readability” score of Wikipedia entries before and after student edits to evaluate students’ ability to apply lessons provided on health literacy and communication skills [11]. Badgett evaluated student edits based on the publication types of the sources (e.g., systematic reviews) the students used to to support their edits [14].

## Discussion

Our findings indicate that Wikipedia editing courses introduce students to EBM skills and provide opportunities to apply these skills. Currently many learners initially encounter EBM in preclinical training [52,53], which can make it difficult to appreciate its relevance to their future clinician roles [13,54]. In contrast, the authentic experience of editing pages to be viewed by potentially thousands of readers is an intrinsically motivating approach to EBM [13,55]. However, a minority of courses explicitly referenced EBM, while the vast majority taught one or more EBM skills, speaking to a missed opportunity to make a direct link. Moreover, instructors should consider longitudinally assessing the integration of EBM in HPE such that learners’ knowledge and behaviors are evaluated not only following the Wikipedia course, but also when students engage in clinical practice.

Editing Wikipedia’s medical content requires editors to first interpret biomedical literature and second translate it into plain language interpretable across a spectrum of health literacy levels. These communication skills are similar to those that health professionals must use when sharing information with patients, skills which when effectively deployed have been shown to impact medical outcomes, safety, and patient satisfaction [56]. Several courses embedded health communication content. One course evaluated students’ experience in attempting to simplify their language [11]. Future instructors might draw upon these courses and their targeted efforts on health literacy to further develop HPE students’ communication skills, which have been documented as deficient upon graduation [57,58].

In contributing to the formulation and sourcing of Wikipedia entries, HPE students are learning about medical epistemology (how health knowledge is assembled, verified, and effectively communicated). This may guide Wikipedia use in their own practice and better position them to support patients searching and interpreting online medical information. In this process, students attain a greater degree of learner autonomy by gaining an ability to critically assess Wikipedia’s strengths and limits, with a new appreciation for the value of its sources. Moreover, students may gain an ability to rectify and enhance this public source as they have the potential for occasional edits and corrections of Wikipedia. Future directions include examining the extent to which the students’ experiences with Wikipedia have contributed to their learner autonomy, especially around delving into Wikipedia’s source materials, their guidance of patient’s use of Wikipedia, and their contributions to it. A longitudinal study examining the effect different educational approaches have on editor and instructor retention (students and professors who continue to improve Wikipedia or continue to teach new course iterations) is also warranted.

Inherent in these courses is the empowering and positioning of students as critics and moderators of knowledge. Indeed, all courses culminated in students critically appraising Wikipedia topics. However, there was variation in how much agency the students received toward that goal. For example, while many courses included peer-evaluation activities, which allowed for epistemic agency and shared evaluation [59-61], only half offered students opportunities to select the article they edited. Wikipedia offers great potential to instructors seeking methods of engaging learner autonomy in their courses and to medical practitioners who take Wikipedia courses to more readily participate in current medical discourse and applying current medical knowledge in their clinical practice.

## Limitations

While we systematically searched the literature and WikiEdu’s dashboards, we possibly missed initiatives, including those that may not have used a WikiEdu dashboard. While the dashboards provided substantial course information, we were unable to capture some materials that were presented as external links nor identify instructor professions. We were also unable to extract information on the quality of the final article edits, including how many met Wikipedia’s guidelines.

We believe we identified the most robust sources of information available on these courses. However, we feel instructors would be challenged to adopt a course based on the identified dashboard or articles alone. We recommend course instructors consider making their materials freely available perhaps by crafting MedEdPORTAL articles or sharing resources via FOAMed, which allow for sharing of detailed curricular materials and would enable instructors to readily examine available materials and make informed judgment on implementing those they deem appropriate.

### Future Directions

Beyond using Wikipedia as a teaching platform and learning tool, these initiatives aimed to improve the quality of Wikipedia’s medical pages many of which are heavily accessed suggesting an impact on public health. Thus, ensuring edits are based on high-quality evidence presented in an unbiased neutral manner is a critical outcome for these courses. We identified a single educational initiative that included a final step of an instructor or expert verifying and adjusting the final product to ensure that the Wikipedia articles were left with high quality contributions [11]. Our results indicate that some instructors included proactive measures to moderate the quality of the final edits, such as encouraging students to interact with and work with Wikipedia’s medical editors, drafting contributions in My Sandbox, assessing contributions for readability and clear language, and peer reviewing suggested edits. Increased implementation of these measures, along with a final review of the live Wikipedia article following the editing initiative by a content expert, can help ensure that students are properly interpreting Wikipedia’s guideline for reliable medical sources. This focus on quality will also improve student experiences and interactions with the Wikipedia volunteer community.

What appears to be missing from the use of Wikipedia editing in HPE is systematic forms of assessment and evaluation to substantiate both the gains in learning and in the quality of Wikipedia entries. Through the Wiki Edu dashboard, there is already a common set of measures for words and references added among the HPE courses. To this, common strategies and instruments could be established among the instructors to establish a body of finding across different settings and cohorts, not so much as a means of grading the students, but as a way of comparing the effectiveness of teaching strategies among different populations of students. These might include pre-test and post-test exercises to furnish the students with a means of assessing changes, for example, in (a) how they would approach the use of Wikipedia by professionals and patients; (b) identifying key factors in constructing and communicating medical knowledge; (c) ways of learning about a new topic; and (d) expertise on the topic on which they have contributed to Wikipedia, and (e) means of engaging in evidence-based medical practices. These assessments might also involve an extension of Azzam’s deployment of outside experts [11] to assess a sample of editing changes for their quality, perhaps along dimensions of accuracy, evidence, and clarity. Such an approach, especially with some coordination among initiatives, might add to the evidence-based pedagogical practices behind HPE, which would be consistent with its own educational goals along EBM lines.

## Conclusion

We believe our analysis presents the most complete record of Wikipedia educational initiatives for HPE students. These courses are actively engaging students in constructing EBM materials for the benefit of others, while learning about the nature and communication of medical knowledge. Based on the increasing number of these courses and instructors’ willingness to continue offering them, there are grounds for more systematic assessments of their contributions to student learning and practices.

## Data Availability

The data for this study consists of published articles and online educational dashboards. Complete references and links are included for these resources in the references section.

